# Weakly Supervised Multiple-Instance Learning for Seizure Onset Zone Identification

**DOI:** 10.64898/2026.09.30.26364032

**Authors:** Junan Mao, Anne-Cecile Lesage, Liliana Camarillo Rodriguez, Diosely C Silveira, Yuanyi Zhang, Patrick J Karas, Jiefei Wang

**Affiliations:** Division of Epidemiology and Biostatistics, University of Illinois Chicago, Chicago, Illinois 60612; Departments of Biostatistics & Data Science, University of Texas Medical Branch, Galveston, Texas 77555; Departments of Neurosurgery and University of Texas Medical Branch, Galveston, Texas 77555; Departments of Neurology, University of Texas Medical Branch, Galveston, Texas 77555; Department of Neurosurgery, University of Texas MD Anderson Cancer Center, Houston, Texas 77030

**Keywords:** epilepsy, seizure onset zone, neural fragility, multiple instance learning

## Abstract

Accurate localization of the seizure onset zone (SOZ) is essential for planning resection or ablation in patients with focal drug-resistant epilepsy (DRE). However, the true SOZ cannot be directly observed during clinical evaluation, so precise electrode-level annotations are rarely available. To address this limitation, we developed a weakly supervised multiple-instance learning (MIL) framework in which labels were constructed from electrode resection status and postsurgical outcome. A shared gated recurrent unit (GRU) with temporal attention encoded each electrode’s peri-onset neural fragility sequence. And attention-based pooling integrated information across variable numbers of electrodes to estimate whether each electrode group contained SOZ-related evidence. The cohort comprised 42 patients with DRE from three datasets, including 29 with successful and 13 with failed postsurgical outcomes. These patients contributed 136 seizure epochs. The cohort was divided at the patient level into training, validation and held-out test sets containing 25, 8 and 9 patients, respectively, with all data from each patient retained in the same subset. Predictions from non-resected regions were evaluated at the seizure-epoch and patient levels to assess postsurgical outcome discrimination for individual seizures and after aggregation across seizures from the same patient, respectively. At the seizure-epoch level, the proposed framework achieved an area under the receiver operating characteristic (ROC) curve (AUC) of 0.828, with a 95% confidence interval (CI) of 0.511-1.000, and performed comparably to Manifold Oblique Random Forests (MORF). At the patient level, the framework achieved an AUC of 0.944, with a 95% CI of 0.677-1.000, and yielded a numerically higher AUC than MORF. However, DeLong’s test did not show a statistically significant difference between the methods, with a P value of 0.398. These findings support the feasibility of extracting clinically meaningful SOZ-related information from electrode-wise neural fragility sequences without precise electrode-level labels. Because the held-out test set included only nine patients and the CIs were wide, these preliminary findings require validation in larger independent cohorts.

## 1 Introduction

DRE affects over 15 million people worldwide and remains associated with substantial morbidity, mortality, and socioeconomic burden. DRE is defined as the failure of adequate trials of two appropriately chosen and tolerated antiseizure medications to achieve sustained seizure freedom.^1^ For patients with focal DRE, surgery or ablation of the SOZ, the brain region critical for seizure generation and propagation, is the most effective therapy, and neuromodulation provides meaningful benefit when resection is not feasible. Across these interventions, clinical success depends on accurately identifying the brain SOZ regions whose removal or modulation maximizes seizure control.

In practice, however, SOZ localization remains challenging because seizures arise from distributed and patient-specific brain networks, while electrophysiological signatures can vary substantially across individuals and recording settings.^2^ A broad range of electrophysiological and network-based approaches has been explored to improve SOZ localization and related clinical assessment. Signal-level biomarkers such as interictal high-frequency oscillations have shown associations with seizure-generating regions, although their detectability and specificity can vary across patients and recording settings.^3^ Other studies have examined seizure-related risk or network-level dynamics using intracranial electroencephalography (iEEG), including seizure likelihood forecasting and network-control-based analyses of seizure evolution.^4,5^ More recently, neural fragility has emerged as an interpretable dynamical biomarker derived from iEEG, with prior studies reporting associations with clinically annotated SOZ labels and postsurgical outcomes.^6,7^

Despite prior progress, supervised learning for predicting SOZ regions remains limited because the true SOZ is not directly observable during clinical evaluation. The ground-truth SOZ regions can only be partially inferred from the resected regions and the postsurgical outcome. Fig. 1 illustrates the assumptions underlying weak-label construction. A successful postsurgical outcome is taken to suggest adequate coverage of seizure-generating tissue, whereas a failed outcome may indicate that relevant tissue remained outside the resected region. These relationships are modelling assumptions rather than electrode-level ground truth. This limitation motivates a weakly supervised learning framework that uses resected regions and the postsurgical outcome to predict SOZ-related evidence without precise electrode-level labels.

**Fig 1.**
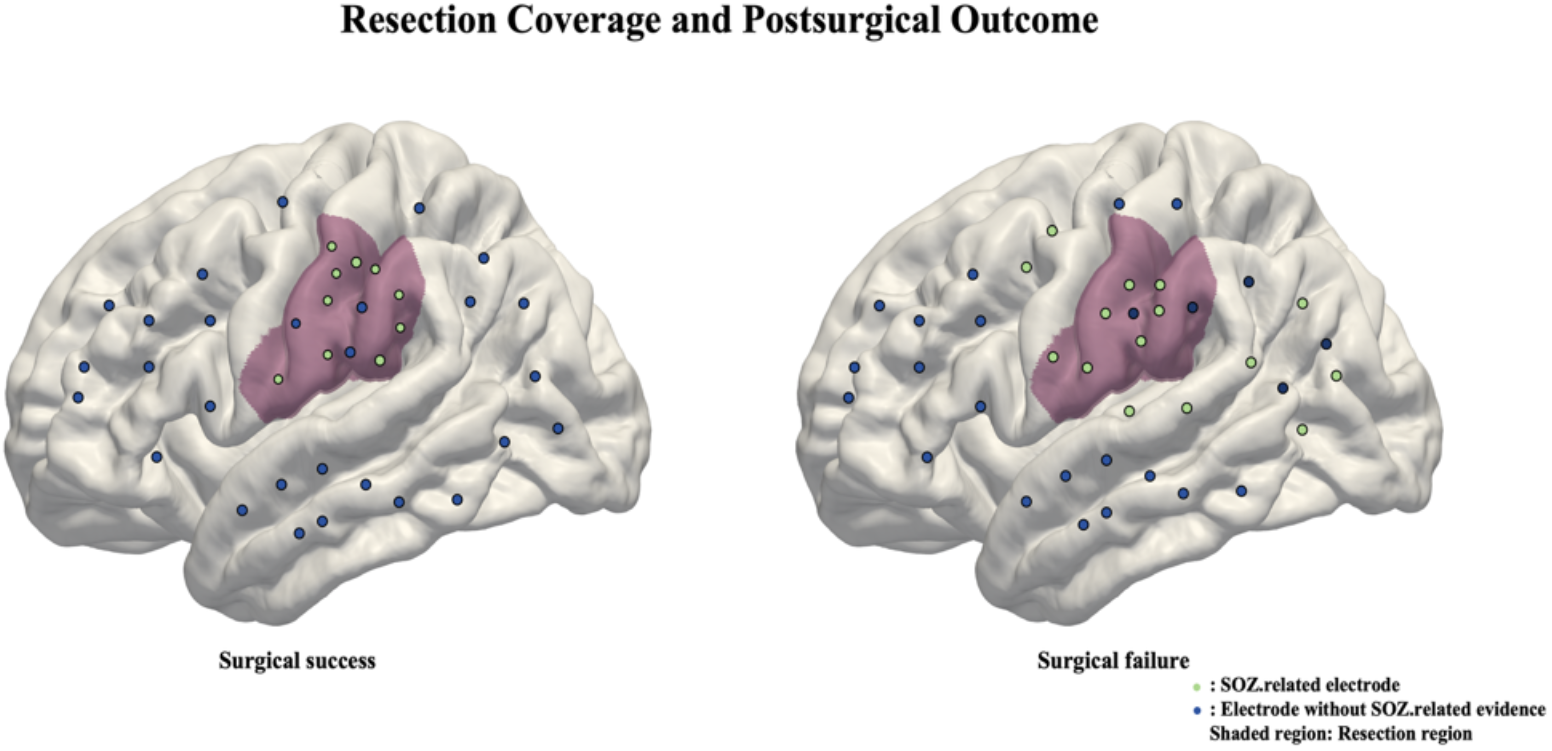
Illustration of resection coverage and postsurgical outcome underlying weak-label construction. The purple shaded area represents the resected region. In the schematic successful case, electrodes with SOZ-related evidence are contained within the resected region, whereas in the schematic failed case, SOZ-related electrodes remain outside the resected region. These scenarios motivate the weak-label rules summarized in Table 1 and represent modelling assumptions rather than electrode-level ground truth.

Neural fragility is a dynamical measure derived from iEEG that reflects how sensitive local brain-network activity is to small perturbations. Changes in neural fragility around seizure onset may therefore provide useful information about seizure-related activity.^6,7^ To use this information without requiring precise electrode-level SOZ labels, we formulated the problem as a weakly supervised MIL framework. For each epoch, electrodes were grouped into bags based on resection status, with electrode-wise fragility sequences treated as instances and bag labels derived from resection status and postsurgical outcome. A shared GRU encoded the temporal evolution of each electrode’s fragility sequence, temporal attention emphasized informative peri-onset time points, and bag-level attention aggregated the electrode representations into a group-level prediction. In this study, we evaluated whether this framework could identify SOZ-related evidence from electrode groups in patients with DRE. Performance was assessed using patient-wise training, validation, and held-out test sets at the bag, epoch, and patient levels, with MORF included as a baseline comparator.^8^

**Table 1.**
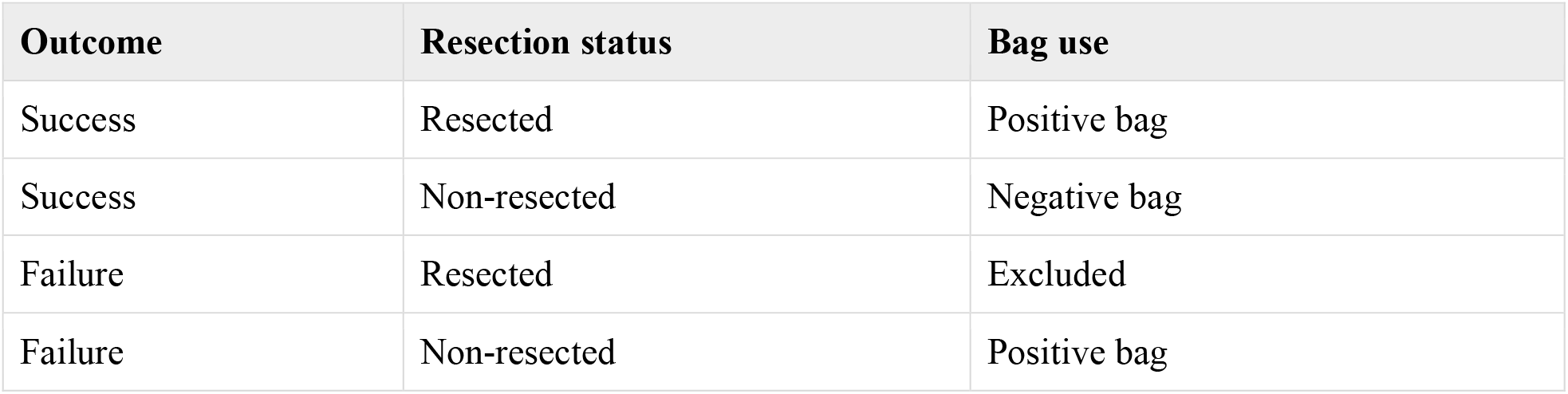
Weak-label rule for MIL bag construction.

| Outcome | Resection status | Bag use |
| --- | --- | --- |
| Success | Resected | Positive bag |
| Success | Non-resected | Negative bag |
| Failure | Resected | Excluded |
| Failure | Non-resected | Positive bag |

## 2 Methods

### 2.1 Data

#### 2.1.1 Data Acquisition

We used Epoch datasets from the University of Texas Medical Branch (UTMB), the Hospital of the University of Pennsylvania (HUP), and the Fragility Multi-Center Retrospective Study. The retrospective UTMB dataset included patients treated from 2022 to 2026. The HUP iEEG Epilepsy Dataset (ds004100) and the Epilepsy-iEEG-Multicenter-Dataset (ds003029) are publicly available through OpenNeuro, with full dataset links provided in the Data Availability section. All three datasets contained data from patients undergoing clinical evaluation for DRE, including iEEG recordings, electrode resection status, and postsurgical outcomes. We downloaded the public Epoch datasets using EpochDownloader in the Epoch package in R.^9^

For a given epoch, the raw iEEG data were stored as a matrix, with electrodes indexed by rows and time samples indexed by columns. Each epoch was associated with electrode-level resection status and patient-level postsurgical outcome information. The resection status identified which electrodes were located within the surgically treated region, whereas the postsurgical outcome provided patient-level clinical evidence for weak-label construction. Across all three datasets, electrode resection status, rather than clinically identified SOZ status, was used to construct the weak labels. No mapping between clinical SOZ annotations and resection labels was performed.

#### 2.1.2 iEEG Preprocessing and Fragility Sequence Generation

The iEEG recordings were reviewed by Dr. Silveira to validate seizure onset times and to identify bad channels for exclusion. The reviewed recordings were then preprocessed in RAVE, including notch filtering and common average referencing. The preprocessed iEEG signals were resampled to a sampling rate of 1000 Hz. We computed electrode-wise neural fragility sequences for each epoch using the EZFragility package, following the previously described neural fragility framework. Consistent with the prior neural fragility study, we estimated neural fragility by fitting linear time-varying models within sequential 250 ms windows and advancing the window by 125 ms, yielding fragility sequences with a temporal spacing of 0.125 s.

After preprocessing and fragility computation, we retained fragility values from 30 seconds before seizure onset to 29.75 seconds after seizure onset for each epoch. With the 0.125 s temporal spacing defined above, this window yielded a common sequence length of *T* = 479 for each electrode.

#### 2.1.3 Epoch-level Normalization

To reduce scale differences across epochs, we performed a z-score standardization for each epoch as follows:

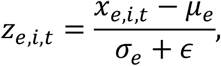

where *X*_*e*_ denote the fragility matrix for epoch *e*, and *x*_*e,i,t*_ denote the fragility value of electrode *i* at time point *t*. The mean μ_*e*_ and standard deviation σ_*e*_ were computed over all entries of *X*_*e*_. ϵ is a small constant added to ensure numerical stability. In this study, we set ϵ = 10 ^− 8^. After normalization, each epoch was represented as a single-channel fragility tensor

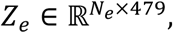

where *N*_*e*_ denotes the number of valid electrodes in epoch *e*.

#### 2.1.4 Machine Learning Data Preparation

After epoch-level normalization, we prepared the data for MIL by constructing electrode bags from each normalized epoch. Each bag contained a subset of electrodes from one epoch. Each electrode-wise fragility sequence was treated as an instance, and the label was assigned at the bag level.

Bag labels were assigned from resection status and postsurgical outcome, as summarized in Table 1. These labels were used as indirect supervision because the true SOZ cannot be directly observed at the electrode level.

The size of non-resected bags in successful cases was sometimes much larger than that of resected bags. To reduce bag-size imbalance, we split large negative bags according to the number of electrodes they contained. A successful non-resected bag with at most 130 electrodes was kept as one bag; a bag with more than 130 and at most 190 electrodes was split into two negative sub-bags; and a bag with more than 190 electrodes was split into three negative sub-bags. In each case, electrodes were divided as evenly as possible so that the resulting bags had similar electrode counts. All split sub-bags inherited the negative label. The output of this step was an outcome-aware collection of weakly labeled bags, each containing a variable number of normalized electrode-wise fragility sequences.

### 2.2 Electrode Encoding and Temporal Attention

This subsection describes the shared electrode encoder shown in Fig. 2. To keep electrode-level encoding consistent across bags while allowing each bag to contain a different number of electrodes, we use a shared electrode encoder for all electrode sequences. For a bag *B*, this encoder processes its *n* electrode-wise normalized fragility sequences separately. All electrode sequences are processed with the same GRU and temporal-attention parameters.

**Fig 2.**
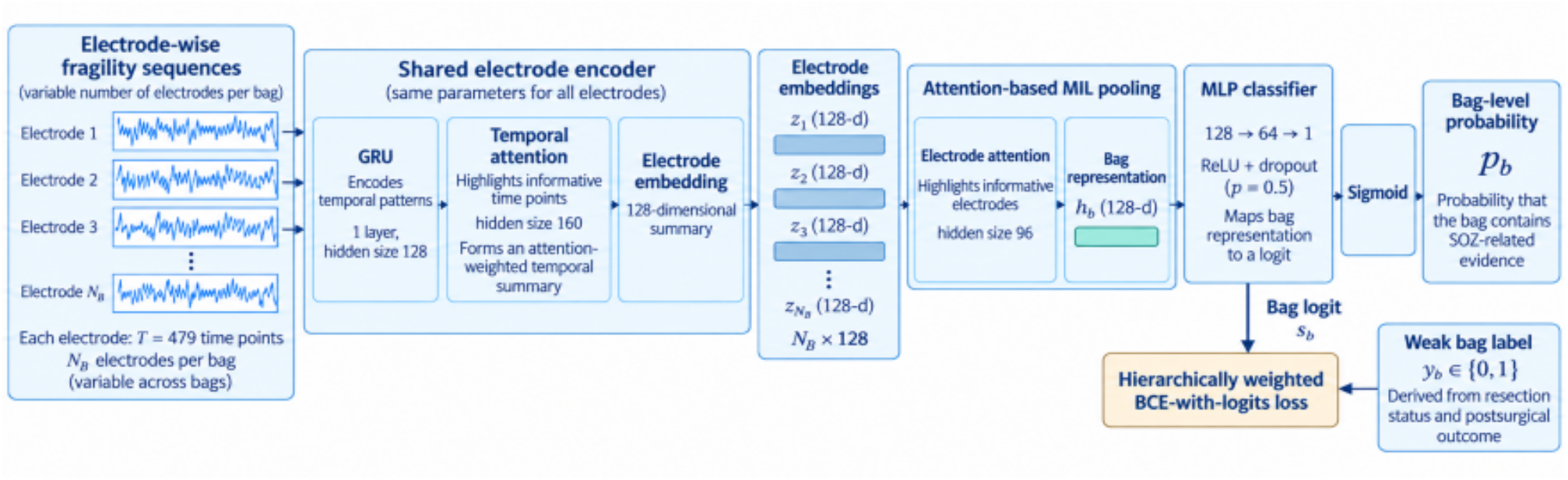
Architecture of the weakly supervised GRU–MIL framework. Each bag contains a variable number of normalized electrode-wise fragility sequences, with 479 time points per sequence. Each electrode sequence is processed independently by a shared one-layer GRU (hidden size 128), followed by temporal attention and a linear projection to obtain a 128-dimensional electrode embedding. Attention-based MIL pooling assigns normalized weights across electrodes and combines the electrode embeddings into a 128-dimensional bag representation. The pooled representation is passed through an MLP classifier with a 64-dimensional hidden layer, ReLU activation and dropout of 0.5 to produce a bag-level logit. A sigmoid function converts the logit to the predicted probability that the bag contains SOZ-related evidence. During training, the bag-level logit and weak bag label are used to compute a hierarchically weighted BCE-with-logits loss.

For the *i*-th electrode sequence in bag *B*, the input is a normalized fragility sequence of length *T* = 479. This sequence is passed through a one-layer GRU with hidden size 128. The GRU produces a hidden state ***h***_*i,t*_ ∈ ℝ^128^ at each time point, where *t* = 1, …, *T*. This step encodes how the fragility signal evolves over time within each electrode, rather than reducing the sequence to manually defined summary features.

Because SOZ-related information may be concentrated in specific portions of the peri-onset sequence, we apply temporal attention over the GRU hidden states for each electrode sequence. Let α_*i,t*_ denote the temporal attention weight assigned to time point *t* for the *i*-th electrode sequence in bag *B*. These attention weights are normalized over the temporal dimension and satisfy

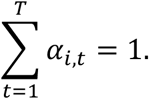

Using these weights, the electrode-level temporal summary is computed as

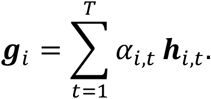

The summary ***g***_*i*_ is then passed through a linear layer and a rectified linear unit (ReLU) activation to obtain a 128-dimensional electrode embedding ***r***_*i*_. The linear layer maps the attended temporal summary into the electrode-embedding space used for bag-level pooling, while the ReLU activation adds a simple nonlinearity to the representation. These electrode embeddings serve as the input to the subsequent bag-level aggregation step.

### 2.3 Bag-Level MIL Pooling and Classification

After the electrode encoder, each bag *B*_0_ is represented by *n*_0_ electrode embeddings, denoted by 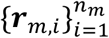. These embeddings are aggregated by the bag-level attention pooling module shown in Fig. 2. Because a bag is an unordered set of electrode sequences, the aggregation function should not depend on the order of electrodes. At the same time, different electrodes within the same bag may contribute differently to the bag-level prediction. Therefore, we use attention-based MIL pooling to assign weights to electrode embeddings according to their contribution to the bag-level prediction.^10^

For the *i*-th electrode embedding ***r***_*i*_ in bag *B*, the attention scoring network first produces an unnormalized attention score. These scores are normalized by a softmax over the *n*_0_ electrodes in the same bag, yielding bag-level attention weights β_*i*_, with

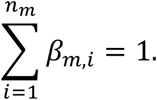

In the implementation, the attention scoring network is a small feed-forward network with hidden dimension 96.

The bag-level representation ***v*** is then passed into a multilayer perceptron (MLP) classifier. The classifier outputs a logit, which is converted into a bag-level probability using the sigmoid function. Let this probability be denoted by *p*, defined as

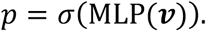

Here, *p* represents the predicted probability that bag *B* contains SOZ-related evidence. In the implementation, the classifier consists of a linear layer from 128 to 64 dimensions, a ReLU activation, a dropout layer with probability 0.5, and a final linear layer from 64 to 1 dimension. The bag-level attention weights β_*i*_ also provide an electrode-level summary of which electrode sequences contribute more strongly to the bag-level prediction.

### 2.4 Training and Evaluation

We evaluated the model using a patient-wise split into training, validation, and held-out test sets. Patients were stratified by postsurgical outcome and assigned to the training, validation, and held-out test sets with a ratio of 60%, 20%, and 20%. The split was performed at the patient level, so all epochs and derived bags from the same patient were assigned to only one subset. This design prevented information leakage across patients during model training, validation, and final testing.

The model was trained to predict the weak binary label assigned to each bag. For each bag *B*_*m*_, the classifier produced a probability *p*_*m*_, representing the predicted probability that the bag contained SOZ-related evidence. We used a sample-weighted binary cross-entropy (BCE) objective at the bag level to penalize disagreement between the predicted probability *p*_*m*_ and the weak bag label *Y*_*m*_. Bag weights were normalized within each patient to reduce the influence of patients contributing different numbers of epochs or derived bags. The checkpoint with the highest epoch-level ROC-AUC on the validation set was selected as the best model and was subsequently evaluated on the held-out test set.

Model performance was evaluated at the bag, epoch, and patient levels. Bag-level ROC-AUC evaluated the primary MIL objective by comparing the predicted bag probability *p*_*m*_ with the weak bag label *Y*_*m*_. Epoch-level ROC-AUC was computed from non-resected candidate bags. For each epoch, the maximum predicted probability among its non-resected candidate bags was used as the epoch-level score. Because multiple epochs from the same patient may be correlated, each epoch from patient *p* was assigned a weight of 1/*n*_2_, where *n*_2_ denotes the number of epochs contributed by that patient. Failed epochs were treated as the positive class, and successful epochs were treated as the negative class. The epoch-level ROC-AUC was then calculated using these weights. Patient-level ROC-AUC was computed by aggregating non-resected candidate bag probabilities across all epochs from the same patient.

### 2.5 Comparison Method

We used MORF as a structured non-deep comparator for the proposed MIL framework.^8^ It was selected because the MORF study included epilepsy surgical outcome prediction using neural fragility.^6^ For each valid epoch, fragility values were standardized using a single mean and standard deviation calculated over all electrodes and time points. The electrodes were then divided into resected and non-resected groups. At each time point, we calculated ten quantiles, from 0.1 to 1.0 in increments of 0.1, separately for the resected and non-resected electrode groups. These summaries formed a 20 × 479 quantile-by-time matrix for each epoch, where the first 10 rows represented the resected-electrode quantiles and the next 10 rows represented the non-resected-electrode quantiles. The 479 columns represented time points from −30 s to +29.75 s relative to seizure onset, using the same 0.125 s temporal spacing as the MIL analysis. Each matrix was treated as a two-dimensional input for MORF, preserving the temporal and quantile structure of the fragility representation. Each valid epoch was treated as an independent training sample and inherited the surgical outcome label of its patient. The prediction target for MORF was postsurgical outcome, with surgical failure treated as the positive class. For consistency, MORF was evaluated using the same fixed patient-wise training, validation, and held-out test split as the proposed model. For evaluation, epoch-level ROC-AUC was calculated with patient-balanced epoch weights, and patient-level scores were obtained by averaging epoch-level failure probabilities within each patient.

### 2.6 Statistical Analysis

Model discrimination was evaluated using AUC. Patient-level uncertainty was assessed using 1000 non-parametric bootstrap replicates generated by resampling patients with replacement. Bootstrap samples containing only one outcome class were excluded from the analysis. The 2.5th and 97.5th percentiles of the valid bootstrap AUC distribution were used to construct 95% CIs. Patient-level AUCs of MIL and MORF were compared using DeLong’s non-parametric test for correlated ROC curves.^11^ Because both methods were evaluated on the same held-out patients, their AUC estimates were correlated. The test accounts for this correlation when estimating the variance of the AUC difference and tests whether that difference is zero. All statistical tests were two-sided, with P < 0.05 considered statistically significant.

## 3 Results

### 3.1 Descriptive Statistics

The study cohort included 42 patients with DRE, including 29 patients with successful postsurgical outcomes and 13 patients with failed outcomes. The cohort contained 136 seizure epochs, with more epochs from successful cases than from failed cases. Patients were split into training, validation, and held-out test sets at the patient level, and each subset contained both successful and failed cases. This split kept all epochs from the same patient within a single subset. MIL bag construction produced 260 weakly labeled bags from the normalized fragility epochs. The validation and held-out test sets had similar numbers of bags and comparable positive-negative bag distributions. Detailed patient, epoch, and bag counts for each split are reported in Table 2.

**Table 2.** Summary of patient, epoch, and bag counts across data splits. Patient counts are reported as total success and failure cases. Bag counts are reported as total positive and negative bags.

| Split | Patients | Epochs | Bags |
| --- | --- | --- | --- |
| Training | 25 (17 success / 8 failure) | 77 | 144 (77 + / 67 -) |
| Validation | 8 (6 success / 2 failure) | 27 | 56 (27 + / 29 -) |
| Test | 9 (6 success / 3 failure) | 32 | 60 (31 + / 29 -) |
| Total | 42 (29 success / 13 failure) | 136 | 260 (135 + / 125 -) |

### 3.2 Bag-Level Performance

Fig. 3 presents the bag-level classification performance of the proposed model on the held-out test set. The test set contained 60 bags, including 31 positive bags and 29 negative bags. The model achieved a bag-level ROC-AUC of 0.892, indicating good discrimination between positive and negative bags.

**Fig 3.**
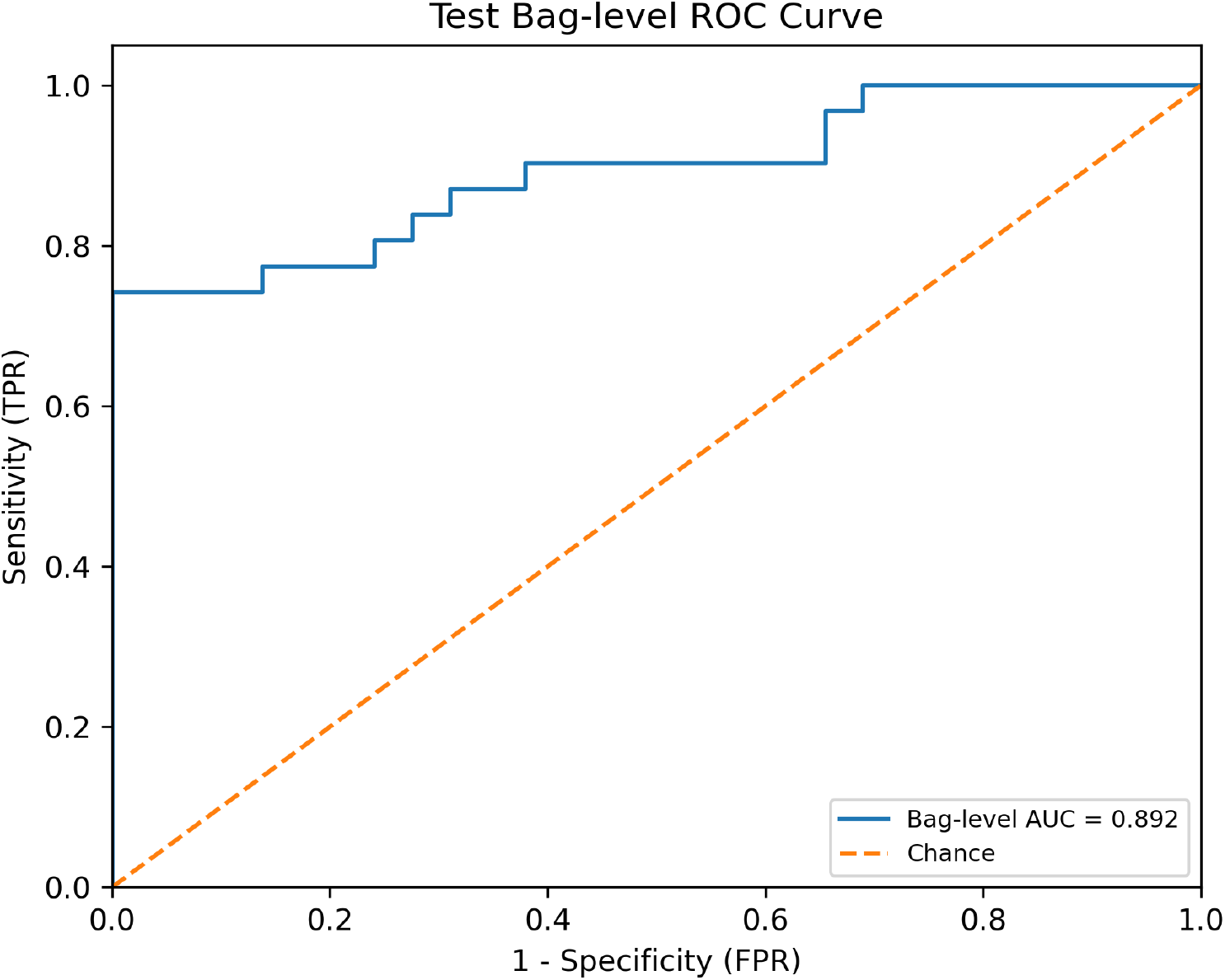
Bag-level ROC performance on the held-out test set. The ROC curve summarizes discrimination of weak bag labels for 60 bags from nine held-out patients, including 31 positive and 29 negative bags. The unweighted bag-level AUC was 0.892. The vertical and horizontal axes show the true positive rate (TPR) and false positive rate (FPR), respectively. The dashed diagonal line indicates chance-level discrimination.

The ROC curve remained above the chance line across most operating points, suggesting that the model generally assigned higher scores to positive bags than to negative bags. Because bag-level classification is the primary objective of the MIL framework, this result provides direct evidence that the model learned informative bag-level representations from the fragility sequences.

### 3.3 Epoch-Level Performance

Fig. 4A presents the epoch-level classification performance of the proposed MIL model on the held-out test set. The model achieved an epoch-level ROC-AUC of 0.8281, with a bootstrap 95% CI of 0.511 to 1.000. For comparison, MORF achieved an epoch-level ROC-AUC of 0.8419 with a bootstrap 95% CI of 0.540 to 1.000, as shown in Fig. 4B. The two methods showed similar epoch-level performance. Although MORF achieved a slightly higher ROC-AUC, both estimates had wide and overlapping CIs.

**Fig 4.**
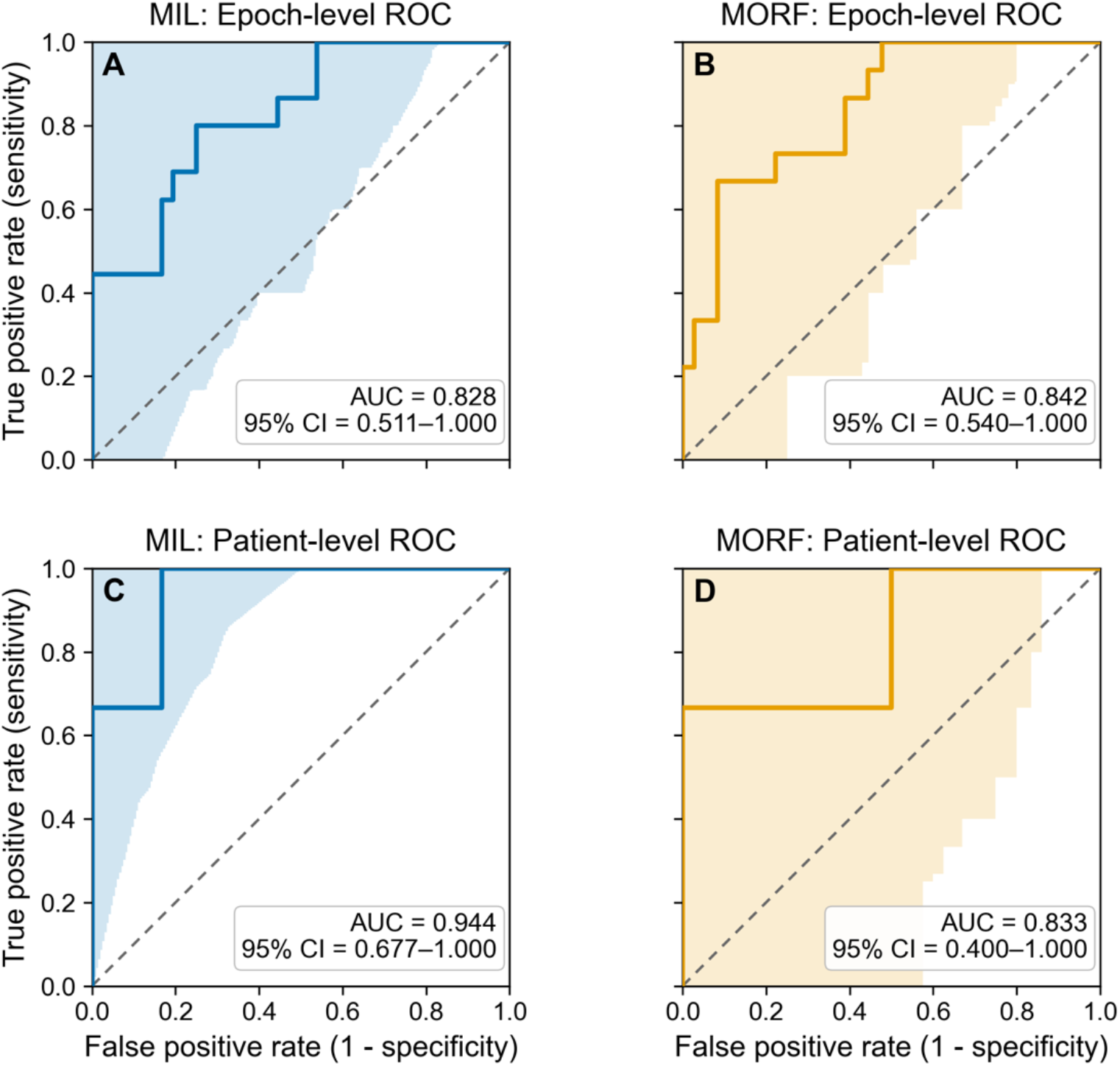
Epoch- and patient-level ROC performance of MIL and MORF on the held-out test set. A, Epoch-level performance of MIL. For each epoch, the maximum predicted probability among non-resected candidate bags was used as the epoch-level score. The patient-weighted AUC was 0.828, with a 95% CI of 0.511 to 1.000. B, Epoch-level performance of MORF. The patient-weighted AUC was 0.842, with a 95% CI of 0.540 to 1.000. C, Patient-level performance of MIL. The patient-level score was defined as the maximum epoch-level score across all epochs from the same patient. The AUC was 0.944, with a 95% CI of 0.677 to 1.000. D, Patient-level performance of MORF. The AUC was 0.833, with a 95% CI of 0.400 to 1.000. The patient-level held-out test set contained nine patients, including six with successful and three with failed postsurgical outcomes, with surgical failure treated as the positive class. DeLong’s test yielded a P value of 0.398 for the patient-level comparison between MIL and MORF. Shaded regions indicate 95% bootstrap confidence bands, and diagonal dashed lines indicate chance-level discrimination.

### 3.4 Patient-Level Prognostic Outcome Performance

Fig. 4C presents the patient-level classification performance of the proposed MIL model on the held-out test set. The model achieved a patient-level ROC-AUC of 0.944, with a bootstrap 95% CI of 0.6767 to 1.000. For comparison, Fig. 4D presents the patient-level performance of MORF on the same held-out test set, with a ROC-AUC of 0.833. Although the proposed MIL model achieved a higher ROC-AUC, DeLong’s test yielded a p-value of 0.398, indicating that the difference between the two methods was not statistically significant.

## 4 Discussion

Our findings suggest that clinically meaningful SOZ-related patterns can be recovered from iEEG fragility under weak supervision. Rather than requiring precise electrode-level annotations, the proposed framework learns from resection status and postsurgical outcome, which provide clinically available but indirect sources of supervision. This formulation aligns the learning task with information routinely recorded in epilepsy surgery cohorts while avoiding the need for manual labeling of brain regions. Taken together, these findings support the feasibility of weakly supervised MIL for extracting SOZ-related information from electrode-wise fragility sequences.

Bag-level performance directly reflects the primary learning objective of the proposed MIL framework. The results suggest that the model could distinguish positive and negative weakly labeled electrode groups using peri-onset fragility patterns. This suggests that electrode-wise temporal encoding and attention-based pooling can extract useful information from electrode groups of different sizes. However, bag-level discrimination should be interpreted as identifying fragility patterns associated with the clinically defined bag labels, rather than as direct evidence of true electrode-level SOZ localization.

Beyond bag-level discrimination, we evaluated whether epoch-level predictions were associated with postsurgical outcome. The MIL model and MORF showed similar epoch-level performance. MORF achieved a slightly higher ROC-AUC, but the wide and overlapping CIs indicate substantial uncertainty in both estimates. In this application, MORF operates on group-level quantile summaries, so its feature-level interpretation is defined over quantile-time features rather than individual electrodes. By contrast, the proposed MIL framework retained electrode-specific representations while achieving performance comparable to MORF, providing a basis for subsequent electrode-level analysis.

At the patient level, the proposed MIL model achieved a higher ROC-AUC than MORF. The bootstrap 95% CI for the MIL ROC-AUC excluded 0.5, indicating statistically significant patient-level discrimination above chance. However, DeLong’s test did not detect a statistically significant difference between the two methods, and both estimates had wide CIs. These findings support above-chance patient-level discrimination by MIL but do not establish that it outperforms MORF.

## Data Availability

All data produced in the present work are contained in the manuscript.

## 5 Limitations

This study has several limitations. First, the cohort was relatively small, particularly at the held-out patient level. The full cohort included 42 patients, whereas the held-out test set contained only nine patients, including three with failed postsurgical outcomes. The limited number of independent patients resulted in wide CIs and reduced the precision of the epoch- and patient-level performance estimates. It also limited the statistical power to compare the proposed MIL framework with MORF and increased the sensitivity of the results to the specific patient-wise data split. Accordingly, the reported findings should be considered preliminary and require confirmation in larger independent cohorts and across repeated patient-wise data partitions.

Second, the available dataset provided only binary postsurgical outcome labels and did not include Engel classifications or quantitative measures of seizure reduction. Consequently, postsurgical outcome was modeled as success or failure, and the analysis could not distinguish different degrees of postoperative improvement. This binary representation may obscure clinically meaningful differences among patients who experienced partial improvement, substantial seizure reduction, or no meaningful improvement. Datasets containing Engel classifications could support ordinal outcome modeling, whereas quantitative measures of seizure-frequency reduction could support continuous outcome analysis. Such information would allow future studies to examine whether model-derived SOZ-related evidence is associated not only with binary surgical outcome but also with the degree of postoperative improvement.

## 6 Data Availability

The data analysed in this study were obtained from the three sources described in Section 2.1. The HUP iEEG Epilepsy Dataset (OpenNeuro ds004100) and the Epilepsy-iEEG-Multicenter-Dataset (OpenNeuro ds003029) are publicly available through OpenNeuro at https://openneuro.org/datasets/ds004100/versions/1.1.3 and https://openneuro.org/datasets/ds003029/versions/1.0.0, respectively. The data were processed using RAVE and the Epoch package in R. Data from the retrospective UTMB cohort are available from the Epoch downloader from the Epoch package in R. Code used for data processing, model training, statistical analysis, and figure generation is available at GitHub: https://github.com/JunanMao/FragilityNet.

